# Medial limbic brain damage selectively impairs affect decoding from normal and whispered voices

**DOI:** 10.1101/2025.05.15.25327699

**Authors:** Marine Bobin, Vivienne Kunz, Lorena C Kegel, Julia Bauer, Hennric Jokeit, Sascha Frühholz

**Author notes:** **Correspondence to:** Marine Bobin & Sascha Frühholz, University of Zurich, Cognitive and Affective Neuroscience Unit, Binzmuhlestrasse 14 (Box 18), 8050 Zürich, Switzerland. and.

## Abstract

The medial temporal lobe (MTL) is crucial for recognizing emotions from various communicative signals, such as voice intonations. It is also a common source of intractable epileptic seizures, and patients undergoing MTL resection are potentially affected by a variety of cognitive and affective deficits depending on the location and size of MTL lesions. This study aimed to identify specific subregions within the MTL where associated lesions lead to deficits in emotional recognition from vocal utterances. This study had a specific emphasis on comparing emotional recognition from normal and from degraded voice signaling, such as in vocal whispering. The latter was assumed to rely on a more extensive processing in MTL regions given the impoverished acoustic signal in whispers. Twenty patients, who underwent unilateral MTL resection targeting the amygdala and hippocampus, and twenty matched healthy controls, were compared using an auditory emotion recognition task from vocal utterances. Participants were asked to recognize and classify the angry, fearful, or neutral tone from whispered or normal vocal utterances. MTL lesion location and spatial extent in patients were determined based on anatomical brain scans, and voxel-based lesion-symptom mapping (VLSM) analysis was applied. The key findings were that patients generally performed similarly to the control group in the emotion recognition task, except in the fear recognition condition. The brain-behavior analysis revealed that an MTL subregion at the junction between the amygdala and hippocampus was associated with deficits in recognizing emotions from whispered voices. Overall, circumscribed MTL lesions can impair emotion recognition from whispered vocalizations to some degree, pointing to the relevance of an integrated amygdala-hippocampal complex for the accurate recognition of emotions from various vocalization types.

## INTRODUCTION

In the healthy brain, the decoding of emotional information from social signals is accomplished by specific integrated neural networks connected with limbic convergence sites or hubs (LeDoux, 2012a; Pessoa, 2017). Prominent among these hubs are the amygdaloid complex and parts of the hippocampal formation, which facilitate connections between several cortico-subcortical networks, especially within the medial temporal lobe (MTL) (Frühholz et al., 2014a). An increasing body of evidence highlights the pivotal roles of these subcortical limbic structures in auditory processing (Camalier et al., 2019; Ceravolo et al., 2021). These structures are integral to decoding specific auditory inputs, such as emotional cues carried in the human voice (Frühholz et al., 2018). Conveyed through various voice intonations, emotional states generate a complex auditory signal that triggers multiple networks via these hubs (Frühholz et al., 2016b). As such, the integrity of these subcortical hubs is particularly important for the listeners to accurately process the perceived information.

The amygdala in particular is a central node in the MTL involved in detecting and processing relevant information from various social stimuli, such as emotions in human voice signals (Dricu and Frühholz, 2016, 2020). Many neuroimaging studies reported significant amygdala activity in response to vocal emotions (Frühholz et al., 2016c), such as the vocal expressions of anger (Frühholz et al., 2012; Frühholz and Grandjean, 2013; Steiner et al., 2022) and fear (Fecteau et al., 2007; Leitman, 2010; Pannese et al., 2016). This connection is further emphasized in clinical contexts, where patients with amygdala damage often display difficulties in recognizing fear and anger in vocal utterances (Scott et al., 1997; Frühholz et al., 2015). Consequently, the amygdala is established as a major hub for deciphering emotional cues in vocal communication.

Located directly posterior to the amygdala, the hippocampus shares substantial anatomical and functional connectivity with this region (Frühholz et al., 2014b). Neuroimaging studies often reported a co-activation of the amygdala and the hippocampus during the processing of auditory emotions (Koelsch et al., 2006; Cheung et al., 2019; Koelsch, 2020). Although the functional contribution of the hippocampus to auditory processing gained increasing knowledge in recent years (Billig et al., 2022), its specific role during the perception of emotional voices remains unclear. It has been suggested that activation in the hippocampus might support the amygdala in emotional decoding (Frühholz et al., 2014b), especially during the decoding of complex emotional voice signals that require enhanced processing efforts (Alba-Ferrara et al., 2011). During such challenging processing conditions, it is hypothesized that the hippocampus retrieves and provides associative, social, and contextual information to support accurate affective decoding (Frühholz et al., 2016c).

A particularly challenging case for emotion decoding arises when dealing with masked or degraded sensory information (Swanborough et al., 2020). In such instances, the hippocampus might play a more pronounced role by providing additional signaling, through pattern completion (Rolls, 2013), likely to rely on memory associations (Horner et al., 2015). Degradation in sensory information can originate from various factors such as contextual elements, signal transmission filtering, or during specific types of vocalizations (Frühholz and Schweinberger, 2021). Whispering represents one such example of vocalization characterized by degraded sensory information (Frühholz et al., 2016a). During whispering, much of the tonal and spectral voice information is missing, while temporal information remains largely preserved. Spectral voice information however seems central for the decoding of many vocal emotions, including vocal fear and anger (Patel et al., 2011). Although the available sensory information in whispered vocalizations, and especially in whispered emotions, is considerably reduced, humans surprisingly still manage to reliably decode emotions from whispered speech (Frühholz et al., 2016a). The recognition rate for emotions conveyed through whispering exceeds expectations given the significant degradation of the voice signal (Deng et al., 2017), suggesting the recruitment of different neural networks relaying compensation mechanisms (Hervais-Adelman et al., 2012). Therefore, we hypothesized that for decoding emotions from acoustic intonations in whispered utterances, the hippocampal formation might support the amygdala by retrieving associations for sensory pattern restoration and completion.

In this study, we explored this potential neural mechanism by testing emotion recognition abilities from whispered and normal utterances, in patients with focal lesions in the MTL, and in matching healthy controls. The temporal lobe is at the origin of various types of epilepsy, characterized by an epileptogenic zone involving the amygdala and hippocampus (Pittau et al., 2012; Suzuki et al., 2019). Besides, temporal lobe epilepsy (TLE) often manifests as an intractable form for which seizures are difficult to control despite appropriate treatment with medications. Patients would then resort to surgical treatment requiring unilateral MTL resection. These MTL resections can vary in spatial extent but primarily focus on the removal of the amygdala and/or hippocampus brain tissue (i.e. selective amygdala-hippocampectomy). Post-surgery assessments in these patients often demonstrate impairments in certain cognitive skills related to memory (Lee et al., 2002; Tanriverdi and Olivier, 2007; Bonelli et al., 2010; Alessio et al., 2013) and affective decoding abilities (Ammerlaan et al., 2008). Testing these patients provides a unique opportunity to examine the contribution of MTL subregions to auditory emotion processing. We accordingly tested whether unilateral damage in MTL subregions of the amygdalar and hippocampal complex would impair the processing of emotional expression in normally phonated (voiced) and especially in whispered expressions of emotions. Both healthy controls and patients performed an emotion recognition and classification task while listening to emotional intonations superimposed on pseudo-speech material, free of any semantic meaning.

## MATERIALS AND METHODS

### Participants

Twenty patients who underwent selective amygdala-hippocampectomy resection as part of the treatment for drug-resistant focal epilepsy gave their consent to take part in this experiment (12 females; age range 19-58y, mean age 37.2y, SD 12.8). Eleven patients had undergone a right-hemispheric MTL resection, and nine patients had undergone a left-hemispheric MTL resection. Criteria for inclusion in the study were: unilateral surgery during adulthood, no psychiatric disorder pre- or post-surgery, no other neurological disorder affecting cognitive abilities, and no post-surgical complication (i.e. no seizures at the time of the recording). The patients were tested on average 1202 days post-surgery, quantified from last the MTL resection to date in case of multiple surgeries. Detailed clinical patient descriptions including the age, sex, time post-surgery, resection side and type are summarized in **Table S1**.

To estimate and compare the performance level of the MTL patients, we also tested twenty age- and sex-matched healthy controls (HC) (12 females, age range 19-56y, mean age 37.6y, SD 12.9) with the identical experimental setup. HC participants did not report any history of psychiatric or neurological disorders and had intact hearing abilities. Moreover, HC had 13 years or less of school education to closely match the educational profile of the patients (**Table S2**).

All participants performed three subtests from the performance IQ section of the abbreviated Wechsler Adult Intelligence Scale (WAIS-III) (Lezak et al., 2004), namely the Picture Completion test, the Block Design test, and Digit Symbol-Coding test. These subtests were used to estimate the IQ score of each participant. All participants were native German speakers or fluent in German, and the tasks were presented in German. Participants gave their written and informed consent prior to the experiment. Ethical approval for this study was obtained by the governmental ethics committee of the Swiss Cantone Zurich.

### Stimulus material and task

Patients and controls performed an emotion recognition experiment with a three-alternative forced choice task on emotions expressed in affective prosodic intonations. The stimulus material consisted of four different speech-like pseudowords with no semantic meaning (“belam”, “namil”, “nolan”, “minad”). Based on these pseudowords, two female and two male speakers (age range 26-29y, mean age 27.5y, SD 1.12) expressed neutral, fearful, or angry emotions in the affective prosody intonation (factor emotional tone). These emotions were expressed either in a voiced (normal intonated voice) or a whispered vocalizations type (factor phonation type) This resulted in a total of 96 different stimuli (mean duration 634ms, SD 173). This stimulus material was identical to that used in one of our previous studies (Frühholz et al., 2016a).

The experiment setup included one practice run prior to the main experiment to familiarize participants with the task. The practice run consisted of twelve stimuli (2 vocalization types * 3 emotional tones, 2 repetitions) that were not used in the main experiment. Participants were asked to respond as fast and as accurately as possible. After the practice run, participants then completed the main experiment that was performed in two different runs. Each run consisted of one block with voiced emotions (48 trials) and one block with whispered emotions (48 trials). The order of voiced and whispered blocks was counterbalanced. Voiced and whispered vocalizations were not presented intermixed in order to prevent any carry-over effects of perceptual processing from voiced vocalizations that could have influenced the subsequent perception of whispered vocalizations.

Stimuli were presented on a laptop on a trial-by-trial basis. Before each stimulus presentation, a fixation cross appeared at the center of the screen to cue the onset of the stimulus (duration 600ms, jitter ± 200ms). The participants were asked to indicate the emotion expressed in the voiced and whispered vocalizations (neutral, anger, fear) with three buttons using the index, middle, and ring fingers of their right hand. Each trial was followed by a silent period (duration 3250ms, jitter ± 250ms) before the next trial started. The order of runs and the response buttons were counterbalanced across participants and matched between the patients and their matching control. Auditory stimuli were presented binaurally through high-quality headphones (Sennheiser HD 280 Pro) at an intensity/loudness level of 70 dB SPL.

### Statistical analysis

Reaction times (RT) and accuracy (number of correct trials relative to the total number of trials) for the emotion recognition task were used to quantify the participants’ ability to classify emotions expressed in voiced and whispered vocalizations. Linear mixed model (LMM) analyses were performed to account for both fixed and random effects, providing detailed information about the variability within and between subjects. LMMs are beneficial for repeated-measures designs, offering a reliable tool for modeling variations across different population samples.

In addition, we conducted post-hoc t-tests to explore specific pairwise differences suggested by the LMM results. These t-tests provide detailed insights into specific condition comparisons, isolating effects of individual conditions which may not be fully captured by the broader LMM analysis. This dual approach ensures a comprehensive analysis of the data, highlighting nuanced differences that are critical for understanding the impact of MTL lesions on vocal emotion recognition. Post-hoc Welsh t-tests were used in cases of non-normal distribution or unequal variance to maintain statistical robustness.

### Multilevel analysis

We conducted LMM analyses to allow for more complex modeling of the data by accounting for random effects (subject-specific variability) in addition to experimental fixed effects. The fixed effects of group (patients, HC), phonation (whispered, voiced), and emotion (neutral, anger, fear) were tested on the dependent variables (RT, accuracy). Contrasts were set to account for comparison between a tested condition against a control condition, for each factor. This setup allowed us to interpret each coefficient of the tested levels in relation to a reference level acting as a baseline. Hence, for the group factor, “patients” was tested against “controls”; for phonation, “whisper” was tested against “voiced”; and for emotional tone, “angry” and “fearful” were tested against “neutral”. Regarding the random effects included in the final model, the selection process was made by comparison between different nested models. The winning model was determined from the chi-squared nested model fit results. Upon comparison between the two models, lower values for the Akaike information criterion (AIC) and the Bayesian information criterion (BIC), combined with a p-value p(>χ^2^)<0.05, determine the better fit to the data. Random effects were estimated with Restricted Maximum Likelihood (REML), and fixed effects were estimated with Maximum Likelihood (ML). All analyses of variance and multilevel analyses were performed on R (version 4.0.2 Release 2020-06-22) through RStudio (version 2023.03.0+386 “Cherry Blossom” Release 2023-03-09 for Windows).

### Neuroimaging acquisition

In order to quantify the extent and location of brain lesions in the patient sample, anatomical scans were acquired from each patient. Anatomical scans were acquired as T1-weighted images on a 3T scanner with a 32-channel head coil (Philips Achieva, Philips Medical Systems). For one patient, the anatomical scan was acquired on Siemens MAGNETOM Skyra 3T scanner (Siemens Healthcare, Erlangen, Germany). Anatomical images were obtained with a T1-weighted 3D MPRAGE sequence with the following parameters: TR 11.1ms, TE 5.1ms, 240 slices, voxel size 0.67mm^3^, matrix size 368x340mm, flip angle 8°.

### Brain lesion quantification

Anatomical scans were preprocessed using the SPM12 toolbox (version 7219; Wellcome Trust Center for Neuroimaging, London, UK; fil.ion.ucl.ac.uk/spm/) running on MATLAB 2021b. All T1-weighted images were reoriented to the AC-PC plane and the voxel size was resampled to 0.5mm^3^. Lesion location and volume were determined by creating binary lesion masks outlining the lesion in 3D. Lesion masks were manually delineated by two independent people on the axial, coronal, and sagittal planes, using the MRIcron software (MRIcro.com).

To ensure a reliable spatial overlap and consistency for the lesion masks provided by the two independent persons, we calculated the Dice Similarity Coefficient (DSC) for the lesion masks of each patient (DSC range: 0.791─0.993, mean 0.887, SD 0.071). These DSC values are at an acceptable level and range according to previous recommendations (DSC > 0.700) (Zijdenbos et al., 1994). Following the reasoning of Zou and colleagues (Zou et al., 2004), a logit transformation (log(DSC)=In[DSC/(1-DSC)]) might be applied to unbound the range of overlap scores with a recommended log(DSC) > 0.847. The log(DSC) values in our study were in an acceptable range of [1.332─5.053]. Lesions masks were finally averaged and normalized to perform group analyses. Normalization was achieved using the Clinical toolbox for SPM12 (github.com/neurolabusc/Clinical) to account for lesioned brain tissue during normalization estimation.

### Voxel-based lesion symptom mapping

To investigate the relationship between lesion extent and location and the task performance in the emotion recognition task, we performed a voxel-based lesion symptom mapping (VLSM) analysis (Bates et al., 2003a). This methodology allows for the examination of the relationship between the tissue damage location and behavioral performance at a voxel-by-voxel level. This is achieved by comparing the behavioral performance of patients with a common area of injury to the performance of a control group to identify crucial brain regions for a given task (Bates et al., 2003b). The relationship between behavioral measures and the location of brain lesion hence helps to understand the importance of a given region for a specific cognitive function, an important complement to brain activation studies (Rorden et al., 2007). For the VLSM analysis in this study, we used the NiiStat software (nitrc.org/projects/niistat) to perform a permutation-based statistical analysis, allowing for hypothesis testing without relying on assumptions about the underlying data distribution.

For this study, we employed a common technique in neuroimaging research known as hemispheric flipping (see also (Hirel et al., 2017; David et al., 2021)), wherein we spatially transformed the left hemisphere lesions of 9 patients onto the right hemisphere, allowing us to combine them with the lesions of the 11 patients that were originally in the right hemisphere. This was done to increase the statistical power for the analysis of MTL lesion effects on the emotion recognition behavior.

### Deficit score for the VLSM analysis

One crucial aspect of the VLSM analysis involves quantifying the behavioral deficit of patients and linking it to the extent of their brain lesion. This deficit score aimed to capture the difference in performance between patients and HC participants in recognizing emotions under the whispered condition, compared to the normally voiced condition. To calculate the deficit, we considered all relevant performance scores: accuracy and RT for each emotion, for both whispered and normally voiced conditions, and for patients and HC.

An intermediate deficit score for whisper performance of each emotion (Δ_WH_) was first calculated as the difference between the whispered (w) and voiced (v) conditions, separately for the accuracy (ACC) and RT measures, including data from patients (Pat) and their matching controls (HC). Hence, for each emotion condition, the following formula was applied:

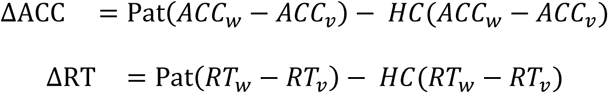

A lower score for accuracy (ΔACC) indicated poorer performance in patients. For RT (ΔRT), a higher score meant that the patients were slower for responding during the task. The general deficit of patients for each emotional tone in the whispered condition (Δ_WH_: Δ_anger_, or Δ_fear_, or Δ_neutral_) was obtained by adding the deficit for whispered voices in accuracy (ΔACC) to the weighed z-scored deficit for whispered voices in RT (ΔRT) of that emotion.

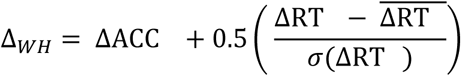

Adding a coefficient to weigh the deficit in response time was done to penalize performance differences, especially in slow and inaccurate responders. The resulting score ensured that the scaling required by NiiStat for VLSM analysis was fulfilled (i.e. NiiStat assumes low values to represent deficits and higher values as good performance). A general patient’s deficit score (GΔ_pat_) was finally calculated with the difference between the deficit in recognizing whispered emotions (Δ_emotion=_Δ_anger_ + Δ_fear_) and the deficit for recognizing whispered neutral tone (Δ_neutral_).

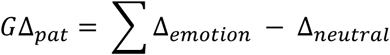

This score, computed for each patient, represented a comprehensive quantification of performance deficits for all emotional whispered trials.

## RESULTS

### Descriptive statistics for the study samples

The patients and HC groups did not significantly differ in age (two-sample t-test; t_38_=-0.086; p=0.932) but had different average years of education (Welch’s t-test; t_23.094_=-3.299; p=0.003) and different IQ scores on average (two-sample t-test; t_38_=-2.480; p=0.018) (**Table 1**). A summary of performance measures related to reaction times (RT) and classification accuracy (percent correct) can be found in **Figure 1** (see also **Figure S1** and **Table S3**).

**Figure 1.**
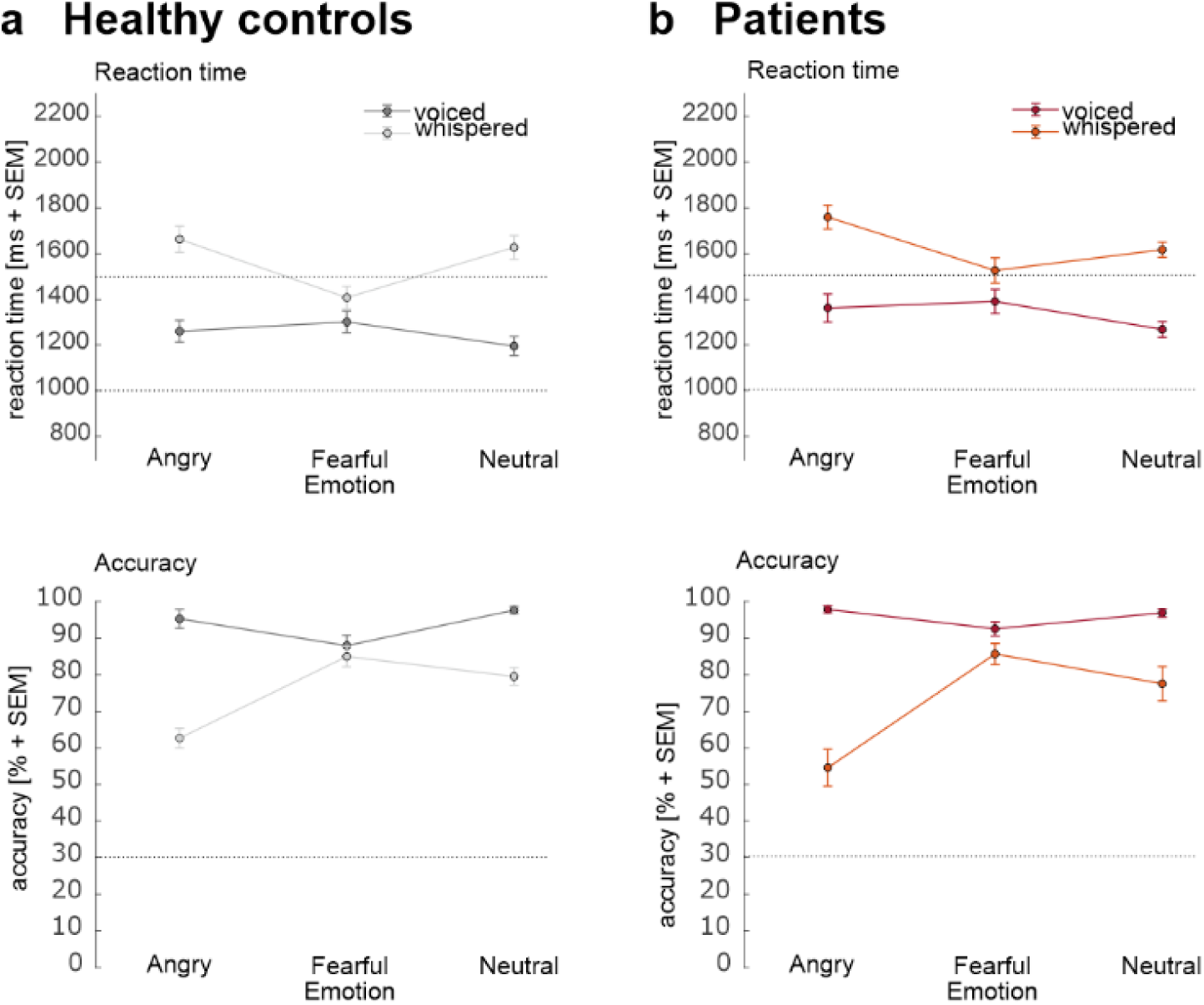
Behavioral performance measures. **(a)** Performance of healthy controls and **(b)** patients in terms of reaction times (upper plots: dashed lines represent the maximal duration of a whispered trial (∼1302 ms) or a voiced trial (∼819 ms)) and performance accuracy (lower plots: dashed line represents chance level of the task).

**Table 1.** Demographical comparison of patients and healthy controls.

| Variables | Healthy controls<br>(n=20) | Patients<br>(n=20) | Two sample t-test<br>statistics | DF | p-value |
| --- | --- | --- | --- | --- | --- |
| Sex (male:female) | 8:12 | 8:12 | - |  | - |
| Age in years (SD) | 37.5 (12.9) | 37.2 (12.8) | t = -0.086 | 38 | 0.932 |
| School education<br>in years (SD) | 12.1 (0.6) | 10.8 (1.7) | t = -3.299 (*Welch*) | 23.094 | 3.125e-3 (***) |
| IQ score (SD) | 108.8 (15.6) | 95.6 (18.0) | t = -2.480 | 38 | 0.018 (*) |
| Signif. Codes p value : '***' p<0.001 ; '**' p<0.01 ; '*' p<0.05 ; '•' p<0.1 |  |  |  |  |  |

Prior to our main analyses as reported below, we first examined the possible influence of lesion lateralization on accuracy and RT to ensure the robustness of our results. We here took into account any potential performance effects due to the side of the lesion in our patient population. A one-way ANOVA was conducted to examine the effect of lateralization on accuracy. The analysis revealed no significant effect of lateralization on performance accuracy (F_1,18_=0.009, p=0.924).

A one-way ANOVA was also conducted to examine the effect of lateralization on RT. The analysis revealed a significant effect of lateralization on RT values (F_1,18_=9.995, p=0.002). To further investigate the specific differences between the lateralization groups, a rmANOVA was conducted with RT as the dependent variable, lesion lateralization as the between-subject factor, and phonation and emotion as the within-subject factors. The analysis revealed a significant main effect of phonation (F_1,18_=102.959, p<0.001, η^2^=0.350) and a significant main effect of emotion (F_2,36_=8.212, p=0.001, η^2^=0.068). However, the main effect of lesion lateralization was not significant (F_1,18_=3.901, p=0.064, η^2^=0.126). Furthermore, there were significant interactions between phonation and emotion (F_2,36_=17.532, p<0.001, η^2^=0.074), but the interactions between lesion lateralization and phonation (F_1,18_=0.290, p=0.597, η^2^=0.002) and lesion lateralization and emotion (F_2,36_=1.092, p=0.347, η^2^=0.010) were not significant. Additionally, the three-way interaction between lesion lateralization, phonation, and emotion was not significant (F_2,36_=0.330, p=0.721, η^2^=0.002).

### LMM model selection

We designed and systematically evaluated several LMMs with various structures of random effects. This comparison between models was crucial to determine the most fitting structure of random effect and capturing the data’s inherent variability and correlation. Our aim was to select the model with the best fit for our data, with the additional goal of keeping the model structure simple.

In the selected linear mixed model (LMM) for accuracy, we included random intercepts and slopes for the effect of phonation for each participant. This structure enabled us to account for individual variability in baseline accuracy as well as in the strength of the relationship between phonation and accuracy performance in the emotion recognition task. The fixed effects in the model included the interaction between groups (patients and controls), phonation, and emotion, allowing us to estimate how these factors jointly influenced accuracy (**Table S2**, for accuracy).

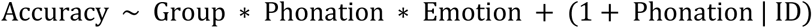

Comparing this model to a simpler model with random-effects accounting for individual differences in the intercepts (i.e. baseline level of accuracy performance) resulted in a better fit for the more complex model (χ^2=^21.219, df=2, p(>χ^2^)<0.001). Adding further complexity in other models and accounting for the random effects of phonation and emotion (χ^2^=9.030, df=7, p(>χ^2^)=0.251) or for the random effects of phonation and group (χ^2^=6.042, df=3, p(>χ^2^)=0.110) to vary across each participant, turned out to be non-significant when compared to the second model (LMM2, cf. Table 5). The AIC and BIC values for this model were the lowest, indicating a better balance between fit and complexity, likely to be an adequate representation of accuracy given the data.

Next, we modelled the RT with a LMM including random intercepts for each participant, and random slopes for the effects of phonation and emotion. This random-effects structure allowed for individual variation across participants in their baseline RT and permits individual differences in how phonation and emotion influence RT (**Table S2**, for RT).

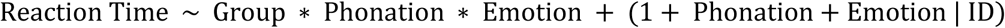

Simpler models were compared, accounting for individual differences in the intercept, and additionally accounting for random slopes for the effect of phonation on RT in each participant (χ^2^=5.772, df=2, p(>χ^2^)=0.056) did not statistically survive the chi-squared comparison to the winning model (χ^2^=22.396, df=7, p(>χ^2^)=2.170e-03). When compared to a model including random slopes for the effects of phonation and group, the comparison still favored the winning model (χ^2^=20.818, df=4, p(>χ^2^)= 3.440e-04). Furthermore, the AIC of the winning model was the lowest of all models, indicating a balanced trade-off between goodness of fit and complexity. The BIC value was however the highest of the proposed models. This criterion is more conservative and penalizes the models with the highest number of parameters. While the winning model is the more complex of the tested models, AIC value and Chi-squared comparison pointed out strong arguments for this choice. Theoretical considerations also played a role in the decision to choose the winning model despite its high BIC value, as the factors that best predicted the variation in individual response times were likely those that varied within the experiment, namely phonation and emotion.

### LMM statistical results for performance measures

The results for the fixed effects from the LMM fit to predict accuracy showed a significant effect of phonation (β=-18.125, SE=3.343, t_137.266_=-5.421, p<0.001), and fearful tone (β=-9.687, SE=2.980, t_160.010_=-3.251, p=1.403e-03). The interactions between phonation and angry tone (β=-14.531, SE=4.214, t_160.010_=-3.448, p<0.001) and between phonation and fearful tone (β=15.156, SE=4.214, t_160.010_=3.596, p<0.001) were also significant (**Table S5a**). No significant effect involving the factor group was observed. **Figure 2** provides a visual representation of the estimates mapped onto the data.

**Figure 2.**
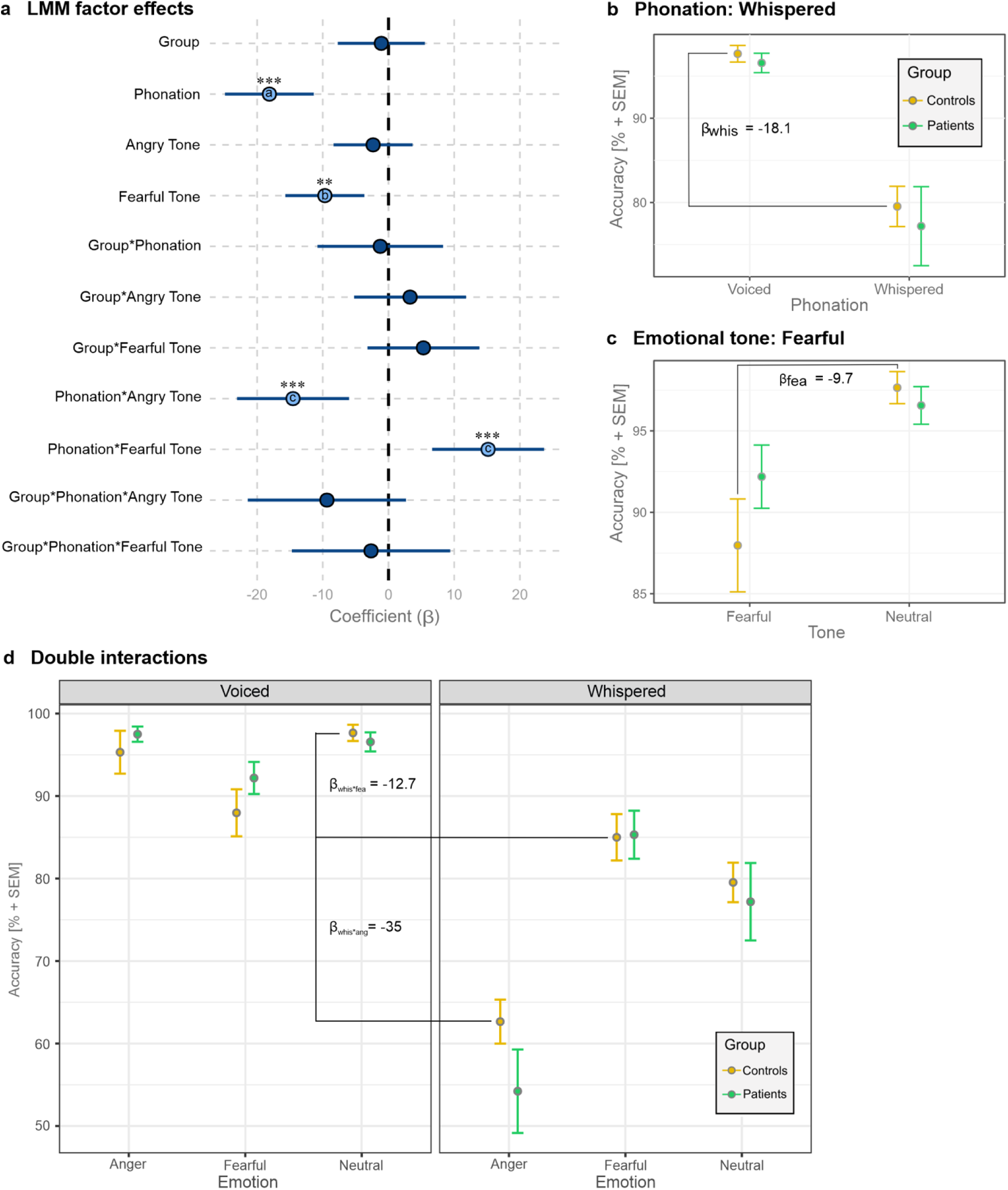
Representation of the estimates obtained with the LMM for performance accuracy. **(a)** Summary of the LMM statistical effects. **(b)** Significant effects were mapped for the whispered condition (vs. voiced condition) and **(c)** for fearful voices (vs. neutral voices). **(d)** Interaction effects between phonation condition and emotional tone. The lines and β values represent the interaction effects, showing how the difference between anger/fear and neutral depends on the phonation condition. These estimate values were calculated by adding each β coefficient composing the interaction: β_whis*ang_ = β_whis_ + β_ang_ + β_whis(ang)_; β_whis*fea_ = β_whis_ + β_fea_ + β_whis(fea)_. The reported coefficients are unstandardized estimates.

In our linear mixed-effects model for the accuracy performance, random effects were estimated to account for variability across subjects (identified by “ID”). The model included random intercepts and random slopes for the effects of phonation within each subject (**Table S5b**). Specifically, the estimated variance for the random intercepts was 0.002, indicating variability in the baseline response time among subjects. The estimated variance for the random slopes associated with phonation (“whispered”) was 0.005, suggesting variation in the phonation effect on RT across subjects. The correlation between the random intercepts and the random slopes for phonation was 0.87, indicating a strong positive relationship between the baseline RT and the slopes of phonation effects within subjects. Additionally, the residual variance, representing variability not accounted for by the random effects, was estimated to be 0.009. The model was fitted using the REML (Restricted Maximum Likelihood) criterion, with a value of -315.7 at convergence. This random-effects structure accounts for individual differences and acknowledges that the data points within each participant are not independent.

In the analysis of the RT data, the fixed effects in the linear mixed-effects model revealed significant main effects and interactions. The fixed effects for phonation were significant (β=431.98, SE=39.58, t_98.16_=10.913, p<0.001), indicating that phonation had a substantial impact on RT. There was also a significant main effect of fearful tone (β=105.82, SE=40.78, t_76.93_=2.595, p=0.011). The effect of angry tone was only close to significance (β=65.09, SE=36.60, t_89.20_=1.778, p=0.079). Additionally, a significant interaction between phonation and the fearful tone condition was observed (β=-325.91, SE=45.70, t_80.02_=-7.132, p<0.001), suggesting that the effect of phonation on RT was moderated by the level of emotion. The main effects and interactions involving the factor group were not significant. Interestingly, the influence of the triple interaction between patient, whisper, and angry tone was close to significance for impacting the task RT (β=113.02, SE=64.63, t_80.02_=1.749, p=0.084) (**Table S6b**). **Figure 3** illustrates the significant coefficients mapped onto the data.

**Figure 3.**
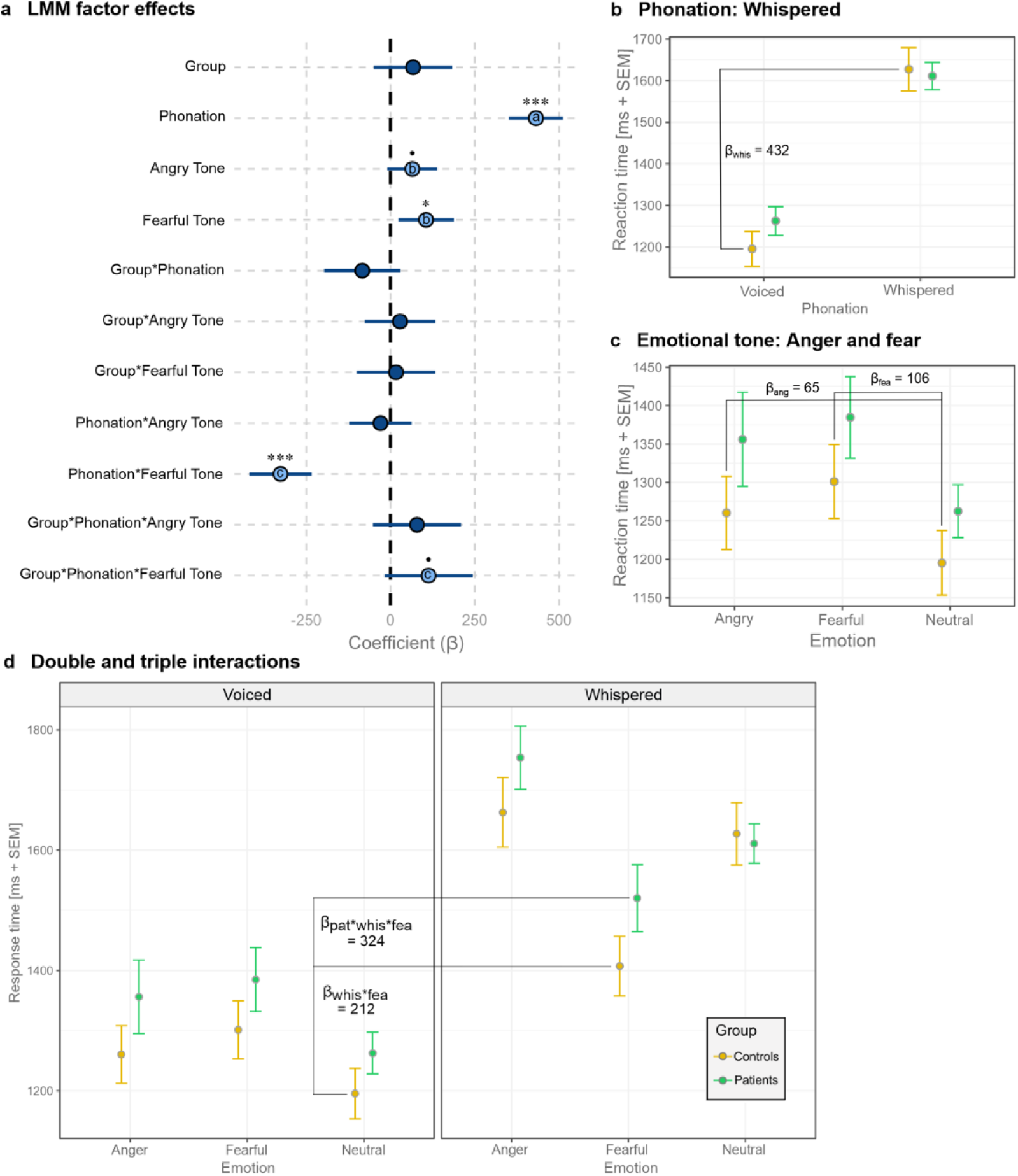
Representation of the estimates provided by the LMM for response time. **(a)** Summary of the LMM statistical effects. **(b)** Significant (or close to significance) effects are mapped onto the data for whispered (vs. voiced) conditions and **(c)** for fearful and angry (vs. neutral) tones. **(d)** The estimates for the double and triple interaction terms were calculated by adding each β coefficient part of the interaction: β_whis*fea_ = β_whis_ + β_fea_ + β_whis(fea)_. For the triple interaction, it involved the coefficients for the main effect of group, whispered, fearful tone, their two-way interaction, and the three-way interaction: β_pat*whis*fea_ = β_pat_ + β_whis_ + β_fea_ + β_pat(whis)_ + β_pat(fea)_ + β_whis(fea)_ + β_pat(whis*fea)._ The reported coefficients are unstandardized estimates.

In addition to fixed effects, the linear mixed-effects model had random effects for individual differences. The random-effects structure included random intercepts and random slopes for phonation, angry tone, and fearful tone across participants (“ID”). The variance components for the random intercepts and slopes were as follows: intercept (variance 23758, SD 154.1), phonation (variance 11002, SD 104.9), angry tone (variance 6225, SD 78.9), and fearful tone (variance 13024, SD 114.1). The residual variance, representing variability not accounted for by the fixed or random effects, was 10991 with a standard deviation of 104.8.

Observations for correlations among the random effects indicated that there was a negative correlation between the intercept and phonation slope (-0.33), and positive correlations between the intercept and emotion slopes (0.47 for anger, 0.21 for fear), and between phonation and emotion slopes (0.48 for anger with whisper, 0.57 for fear with whisper) (**Table S6b**). These random effects indicate that there was significant individual variability in the response times and that the effects of phonation and emotion varied across individuals. This suggests that the impact of phonation and emotion on RT was not uniform across participants, emphasizing the importance of accounting for individual differences in this analysis.

### Pairwise post-hoc comparisons between experimental conditions

After utilizing the LMM to understand the overall behavior and performance differences between the patients and controls, we conducted pairwise comparisons between the whisper and voiced conditions for each emotion (**Table S7**). This dual approach allowed for a comprehensive analysis, combining the broad insights gained from the LMM with the detailed, focused insights from the pairwise comparisons, ensuring a robust and nuanced understanding of the data.

Both accuracy and RT showed significant effects for each comparison (paired t-tests or Wilcoxon signed-rand test, all p<0.001), except for the pairwise comparison for accuracy between whispered and normally voiced fearful tone trials, showing a lower degree of significance (Wilcoxon signed-rank test, p=0.014).

The same pairwise comparisons for the control group showed high significance (paired t-tests or Wilcoxon signed-rand test, all p<0.004). However, the comparison between whispered and normally voiced fearful trials did not reach statistical significance for the control group (Wilcoxon signed-rank test, p=0.109) (**Table S8**).

### Anatomical lesion data

In all patients, the lesion predominantly involved the amygdala, unilaterally, in conjunction with the anterior hippocampus (**Table S1**). The volume of these lesions varied across the patients with a minimum volume of 12’197 voxels, and a maximum of 275’740 voxels (mean volume 84’120.45 voxels, SD 80’853.01). In addition to the amygdala and hippocampus, other parts of the temporal cortex were also affected in a subset of patients. Specifically, the temporal pole, segments of the middle temporal lobe, and the anterior portion of the inferior temporal gyrus were implicated in some cases (**Figure 4**). The diverse localization and extent of these lesions provide a comprehensive view of the variations in medial temporal lobe involvement among the patients studied. Importantly, despite the variability in lesion extent, the surgical interventions were performed with precision to ensure that the lesions were as focal as possible for amygdala-hippocampectomy. This approach aimed to effectively treat drug-refractory epilepsy while preserving as much cerebral tissue as possible.

**Figure 4.**
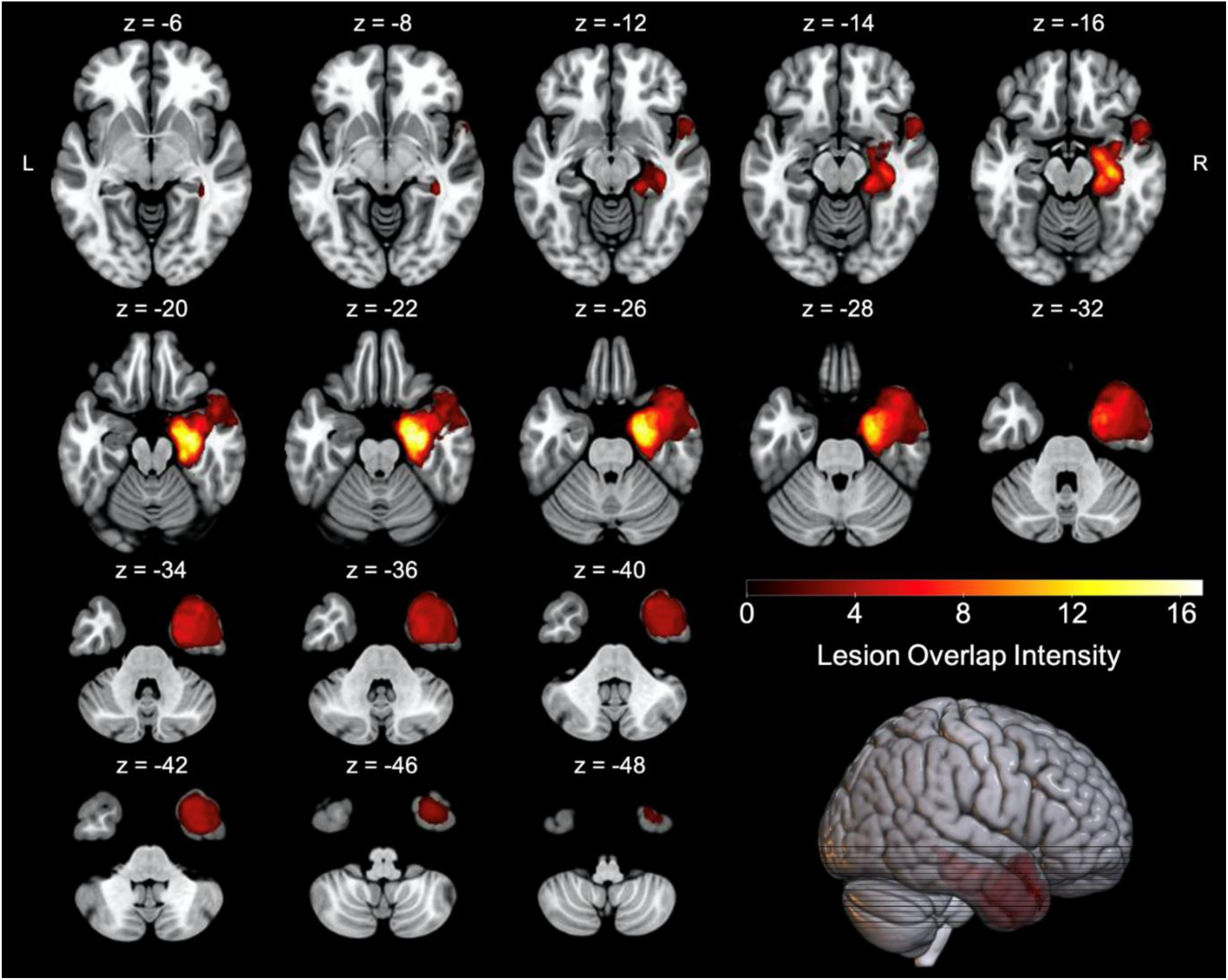
Sum of lesions across patients mapped onto an MNI brain. Slices are displaying an axial view. The scale of color indicates the level of lesion overlap among the patients: the darker red color indicates a weaker overlap (fewer patients), while the lighter color indicates a stronger overlap (more patients).

### Voxel-based lesion-symptom mapping (VLSM)

We investigated which lesion locations were associated with decreased ability to recognize emotions in whispered voices. A VLSM analysis was performed with the composite score expressing the deficit in performance for emotion recognition (vs. neutral) for whispered voices (vs. normal voices). The analysis was focused on MTL lesion in general (ignoring lateralization based on L-R image flipping) (Baldo et al., 2022) and included voxels marked as “lesioned” in at least 10% of the participants (i.e. minimum overlap for at least two patients). We controlled for lesion volume to minimize false positives (Speber and Karnath, 2017). The permutation testing threshold was set to 5000 permutations and corrected to p=0.05 to account for multiple comparisons. The general linear model (GLM) was used for this analysis, including age, gender, onset of epilepsy, and IQ score as covariates to control for their potential influence.

A statistically significant brain-behavior association for deficits in emotional recognition in whispers was observed in a cluster formed of 56 voxels, situated at the junction between the amygdala and the hippocampus. (**Figure 5** and **Table S9**). To further characterize the anatomical composition of this cluster, the MNI (Montreal Neurological Institute) coordinates of the significant voxels were extracted and compared with a probabilistic cytoarchitectonic map (Eickhoff et al., 2005, 2007; Amunts et al., 2007). This analysis revealed that the cluster predominantly consisted of three cytoarchitectonic structures: the subiculum, the superficial amygdala nucleus, and the hippocampal-amygdaloid transition area (HATA) region. In addition to the cytoarchitectonic mapping, macro-anatomical findings located the significant voxels mainly within the hippocampus, parahippocampal gyrus, and amygdala.

**Figure 5.**
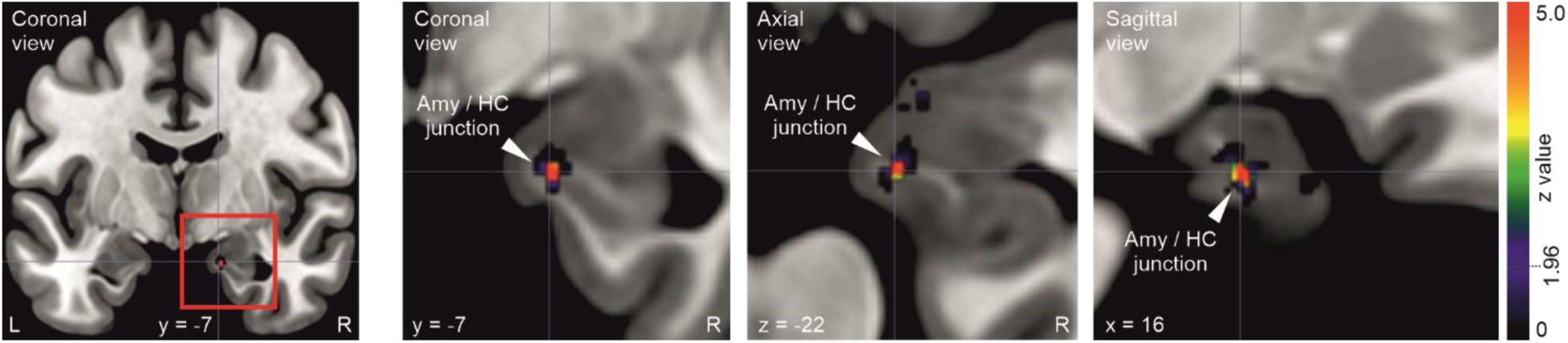
Significant cluster for the VLSM analysis mapped onto a brain in MNI space. The left image shows a whole brain coronal view. The three right images show enlarged views of the area marked with a red box in the left image. Voxel color coding refers to z values (threshold p=0.05, z=1.96).

## DISCUSSION

In this study, we addressed the contribution of the MTL for emotional decoding in vocal utterances through emotion recognition behavior and through voxel-based lesion-behavior mapping. More specifically, our focus was on examining how MTL lesions in the amygdala and hippocampus could impact the ability to recognize emotions in challenging voice perception conditions, such as in whispering. The behavioral component of this study involved an experimental paradigm in which participants were presented with pseudo-words spoken in a normal or whispered voice, and conveying angry, fearful, or neutral emotion in prosodic intonations. High-resolution brain imaging was employed to extract data on the extent of MTL lesions in the patient cohort. Subsequently, we investigated the association between the extent of these lesions and deficits in emotion recognition under challenging speech perception conditions. We found that (a) the performance of patients was negatively impacted by the whispered condition, both in terms of accuracy and response time, although this effect was not different from that observed in the controls; (b) among the different emotional conditions, the performance for fearful vocalizations was the least affected by the phonation type; but patients showed a deficit for fearful whispers which was not observed in the controls, and (c) a lesion at the junction of the amygdala and hippocampus was associated with a more pronounced deficit in recognizing emotions from whispered voices in MTL patients.

### Whispering impairs affective voice recognition

Performance for emotional voice recognition was decreased by the whispered condition across all affective tones and for both groups. For comparison, Frühholz and colleagues (Frühholz et al., 2016a), using the same experiment and stimuli set, did not find any significant effect on performance across the factors of phonation, emotion, or their interaction. Although accuracy was affected by the phonation type, it still remained above a 90% success rate. The difference in age among the participants in the present study could explain this disparity, as it is well-documented that voice signal perception progressively declines across the adult lifespan, especially when in challenging contexts that increase the cognitive load (Goossens et al., 2017). Age-related changes in voice emotion recognition have been observed, and these declines have been shown to occur independently of age-related hearing loss (Christensen et al., 2019). These alterations over time can be attributed to deficits in temporal processing, but also to changes in the neural encoding of voice-related acoustic amplitude modulations occurring with age (Goossens et al., 2018; Regev et al., 2023). Temporal processing of the vocalization envelope and sensitivity to amplitude modulation is critical for the accurate perception of vocal utterances, particularly when discerning emotional nuances (Zhu et al., 2018). Therefore, when voice signals are degraded, the neural mechanisms underlying the processing of temporal envelope variations are especially challenged, which can hinder the recognition of emotional content.

While whispering negatively impacted the overall performance on emotion recognition, the results of the LMM highlighted a differential impact between angry and fearful tones for both groups. Specifically, fearful whispering was more easily recognized compared to neutral whispering, whereas angry whispering was significantly less accurately recognized. Besides this difference in accuracy, angry whispering (vs. neutral whispering) elicited prolonged RT, while fearful (vs. neutral) whispers showed lower RT.

This discrepancy in the efficiency of processing different emotions in whispered speech could be explained by the unlikely combination of the vocal expression of anger in the whispered modality. Typically, vocal anger is characterized by a high intensity, vocalization rate, and pitch, which makes it easily detectable in normal vocalizations (Sobin and Alpert, 1999; Scherer, 2003). However, whispering inherently has impoverished acoustic features, altering the decoding of spectral and temporal information compared to normal voice perception. This requires the auditory system to switch to a more extensive functional network for processing information in whispered vocalizations (Frühholz et al., 2016a). We hypothesize that the cognitive load triggered by whispers compared to normal voices perception results in an efficiency cost, which is reflected in various aspects of performance for the whispered condition. In the present study, this effect is particularly pronounced in the identification of angry voices, but also for neutral voices.

### Fearful condition showed differential effects

The behavioral observations revealed that the fearful tone condition stood out the most. RT was significantly impacted across all tones. But interestingly, RT was more comparable to that of normal voices for fearful whispers. Fearful whispers seemed less affected than other affective tones in terms of processing speed. The results of the LMM analysis indicated a trending three-way interaction, highlighting a subtle difference between groups: the effect of patients (vs. controls) on RT varied depending on whether it was for whispers (vs. normal voice) and fearful (vs. neutral) tone condition. Additionally, pairwise comparisons between whispers and normal voices for patients showed that the accuracy for fearful tone trials was less affected, albeit still influenced, by the difference in phonation. This same comparison was not significant for the controls, meaning that their recognition for fearful voices was not significantly affected by whispers.

An extensive body of literature has documented the evolutionary conservation and significance of fear processing across species (LeDoux, 2012b; Adolphs, 2013). The underlying rationale is that there is an evolutionary advantage in the efficient processing of fear as it signals a potential threat. This has led to the evolution of an integrated nervous system proficient at recognizing and responding to threatening stimuli, ultimately increasing an organism’s chances of survival (Mobbs et al., 2015). While scarce in the auditory domain (Frühholz et al., 2021), studies in the visual modality have highlighted the unique characteristics of fear-related signals in cognitive processing, with some evidence suggesting that fear can be processed even below conscious awareness (Vuilleumier et al., 2002; Bertini and Làdavas, 2021; Làdavas and Bertini, 2021). Further studies hence described the perception of fearful stimuli to engage a specialized neural circuitry, often referred to as the “circuitry of fear”, which involves an interconnected network of brain regions (Tao et al., 2021; Pessoa, 2023). Shaped by both evolutionary pressures and learned experiences, this circuitry has evolved to adapt to varying ecological threats and optimize brain responses to fearful stimuli (LeDoux, 2012a; Hedger et al., 2015). This optimization is believed to confer an advantage in the processing of such emotionally salient signals, facilitating prompt and adaptive reactions to potential threats.

In line with the notion of a ‘fear advantage’ in cerebral processing as previously documented, our current behavioral findings suggest that emotion recognition for fearful tones is more resilient to the acoustic challenges imposed by whispering. For both experimental groups, recognition of fearful tones during whispered speech is less adversely affected, compared to neutral tones (both for accuracy and RT). However, we observed a divergent pattern for fearful tone accuracy between the control group and patients with unilateral lesions in the MTL. While controls did not exhibit a significant difference in accuracy for recognizing fearful tones between the whispered and voiced conditions, patients showed a disparity between these phonation conditions. The ‘fear advantage’ remains an interesting explanation for the current observations, as the accuracy in recognizing whispered fearful tones is still higher compared to other whispered tones. However, the presence of this deficit in patients with MTL lesions, not observed for healthy controls, is consistent with the critical role the MTL plays in emotion processing.

Particularly, all patients in this study had focal lesions involving the amygdala. The role of the amygdala in processing emotional voices has been frequently highlighted in lesioned studies (Frühholz and Staib, 2017). Clinical observations of patients with bilateral amygdala damage reveal an intriguing contrast in that regard: while a case demonstrated impaired perception of intonation patterns, essential to emotional recognition, despite normal hearing (Scott et al., 1997), others showed a sustained ability to evaluate fear in vocal expressions (Anderson and Phelps, 1998; Bach et al., 2013) or intact emotional prosody recognition (Adolphs and Tranel, 1999). Yet, most studies from neuroimaging, electrophysiological, and lesion methodologies converge towards the crucial importance of the amygdala to process verbal emotional content (Frühholz and Grandjean, 2013; Frühholz et al., 2015; Domínguez-Borràs et al., 2019). More specifically, unilateral lesions of the amygdala can result in selective impairments in recognizing fear and surprise in vocal expressions (Dellacherie et al., 2011). Our observations for unilateral MTL-lesioned patients partially aligned with these studies, as the main difference we observed when compared to healthy controls was in the accuracy of fearful whisper recognition.

We observed that patients varied the most in their responses to the fearful whispers. Yet, their performance across experiment conditions was nearly comparable to that of the control group. This is not surprising considering the brain’s plasticity and compensatory mechanisms in the months following surgery (Goldmann and Golby, 2005). Sidhu and colleagues (Sidhu et al., 2016) conducted a longitudinal study and found that in comparison to 3 months post-surgery, the functional engagement of the hippocampus on the opposite side of the brain from the lesion at 12 months translated to an efficient reorganization, regardless of the lesion lateralization. This aligns with the notion that the first year after surgery is a critical period during which significant neural plasticity and behavioral compensation occur (Helmstaedter and Elger, 1998; Stretton et al., 2014).

### Lesion at the junction amygdala-hippocampal formation impairs emotion recognition

Results from the VLSM analysis highlighted a cluster consisting of 56 voxels, situated at the junction between the amygdala and hippocampus, which was associated with the deficit in emotional tone recognition in whispering. Specifically, the significant voxels were located in the subiculum, the superficial (SF) and basolateral (BL) complexes of the amygdala, the hippocampal-amygdaloid transition area (HATA), with partial involvement in the CA1 subfield of the hippocampus, and the entorhinal cortex (EC).

The identified cluster is found within key regions of the brain known to have strong associations with memory and emotional processing. Primarily, a portion of the cluster is situated within the subiculum, part of the parahippocampal gyrus, which bridges the CA1 subfield of the hippocampus and the EC. This region has strong associations with the BL amygdala ^80^ and plays a crucial role in signal integration within the hippocampus by connecting regions that receive inputs from the hippocampus (Chase et al., 2015). Notably, the subiculum has been observed to be particularly active during tasks involving explicit memory and fear perception (Herman and Mueller, 2006). Additionally, the identified cluster encompasses portions of the amygdala, specifically the SF and BL complexes. The SF complex, externally located between the BL and centromedial nuclei of the amygdala, is directly adjacent to the EC and has bidirectional connections with the hippocampal formation (Heimer et al., 1999; Amunts et al., 2005). This amygdalar complex is functionally linked with other limbic regions (Roy et al., 2009) and associated with the detection and processing of emotionally salient stimuli (Kerestes et al., 2017). Concurrently, studies have suggested that the BL complex of the amygdala acts as an integrator of multiple sensory modalities by coordinating higher-level sensory inputs and co-activating with regions such as the associative auditory cortex and the hippocampus (Bzdok et al., 2013). Lastly, the cluster extends to the HATA, which bridges the body of the hippocampus and the caudal amygdala. Characterized by a distinct cytoarchitecture that gradually evolves into the adjacent CA1 field of the hippocampus, the HATA also sends inputs to the medial nucleus of the amygdala (Fudge et al., 2012). This is noteworthy given the association of CA1 with the retrieval of remote episodic memory (Bartsch et al., 2011).

Hence, this cluster’s composition suggests an important role in the observed brain-behavior associations, particularly in recognizing emotional tones during challenging voice perception conditions. The deficit score input into the VLSM analysis indicates an impaired ability to recognize emotional tones (angry, fearful) in whispering for patients (vs. HC). The resulting cluster associated to such a deficit bring together a set of precise subregions that seem to be highly relevant for this ability. The inclusion of the subiculum and amygdala, both essential components of the limbic system, highlights their synergistic role in emotion processing and recognition in the auditory modality. Moreover, the presence of the HATA, which serves as a link for hippocampal-amygdala interactions, highlights the integrated nature of these regions in memory and emotion processing. Previous evidence suggest that the disruption of the HATA is correlated with a diminished capacity for adapting to situational learning and access to emotional memories (Fudge et al., 2012; Wan et al., 2021). Our obervations are in line with this role for HATA.

### Limitations of the study

While our study represents the first to investigate the association between emotion recognition in voice intonations and functional aspects of the MTL in unilateral lesioned patients, it carries several limitations. First, our sample comprises patients with drug-resistant focal epilepsy, a condition known to induce significant individual variability in functional reorganization both pre- and post-surgery, which may impact hippocampal connectivity and cognitive outcomes (Griffin and Tranel, 2007; Doucet et al., 2015; Witt et al., 2015; Elias et al., 2021). This, combined with our relatively small sample size, could obscure relationships between the extent of the lesion and the severity of the behavioral impairment, despite our use of LMMs to account for variability in the behavioral data. Second, a more comprehensive assessment of voice processing abilities, for instance through extensive acoustic testing (Roswandowitz et al., 2018), was not feasible due to the limited accessibility to the clinical population and restrictions on the number of tasks that could be presented. Third, our VLSM methodology, while informative, presents inherent limitations such as inaccurate judgment of the integrity of the brain tissue surrounding the lesion and potential variability in the nature of the damage (Pittau et al., 2012; Blumcke et al., 2017). Additionally, while epilepsy’s specific causes underlying seizures are important, they were considered in this study as a single condition due to the limited sample size. Finally, while our hemispheric flipping approach is commonly used (Meyer et al., 2016; Hirel et al., 2017; Karnath and Rennig, 2017) and helped us focus on the impact of brain damage on a specific cognitive ability, it operates on the assumption of functional and structural symmetry between hemispheres. This could limit the contribution of our findings to the debate around hemispheric specialization for speech and emotional processing. Therefore, future studies with larger sample sizes for each hemisphere would be needed to draw more definitive conclusions.

### Conclusions

The present study investigated deficits in emotion recognition during normal and degraded voice utterances in patients with lesions in the MTL. The voxelwise statistical analysis allowed for the identification of a specific, focal region within the MTL that is strongly associated with emotional recognition during whispered speech. This region is situated at the junction between the amygdala and hippocampus, encompassing subfields that have been shown to be crucial in the processing of emotional stimuli and memory retrieval. While these subregions have not been extensively discussed in the context of emotional voice perception, our observations highlight their involvement in processing emotionally charged auditory stimuli, especially under challenging conditions.

## Data availability

The datasets generated and analyzed during the current study are available from the corresponding author on reasonable request. Upon sharing, they will comply with ethical guidelines as they contain patient-sensitive data. Additionally, the code used for the LMM analysis and VLSM will be made available upon reasonable request.

## Data Availability

The datasets generated and analyzed during the current study are available from the corresponding author on reasonable request. Upon sharing, they will comply with ethical guidelines as they contain patient-sensitive data.

## Acknowledgments

SF was supported by the Swiss National Science Foundation (SNSF PP00P1_157409/1 and PP00P1_183711/1 to SF). SF also received grant support from the Vontobel Foundation (www.vontobel-stiftung.ch; Zurich, Switzerland).

## Authors contributions

MB / VK / SF contributed to designing the experiment, data acquisition, data analysis, and writing the manuscript; LK / JB / HJ contributed to data acquisition.

